# Wastewater monitoring of SARS-CoV-2 RNA at K-12 schools: Comparison to pooled clinical testing data

**DOI:** 10.1101/2022.12.21.22283809

**Authors:** Sooyeol Kim, Alexandria B. Boehm

## Abstract

**Background:** Wastewater measurements of SARS-CoV-2 RNA have been extensively used to supplement clinical data on COVID-19. Most examples in the literature that describe wastewater monitoring for SARS-CoV-2 RNA use samples from wastewater treatment plants and individual buildings that serve as the primary residence of community members. However, wastewater surveillance can be an attractive supplement to clinical testing in K-12 schools where individuals only spend a portion of their time but interact with others in close proximity, increasing risk of potential transmission of disease.

**Methods:** Wastewater samples were collected from two K-12 schools in California and divided into solid and liquid fractions to be processed for detection of SARS-CoV-2. The resulting detection rate in each wastewater fraction was compared to each other and the detection rate in pooled clinical specimens.

**Results:** Most wastewater samples were positive for SARS-CoV-2 RNA when clinical testing was positive (75% for solid samples and 100% for liquid samples). Wastewater samples continued to test positive for SARS-CoV-2 RNA when clinical testing was negative or in absence of clinical testing (83% for both solid and liquid samples), indicating presence of infected individuals in the schools. Wastewater solids had a higher concentration of SARS-CoV-2 than wastewater liquids on an equivalent mass basis by three orders of magnitude.

## Introduction

Wastewater-based epidemiology has been widely used across the world to supplement clinical data on COVID-19. Most research aimed to understand the relationship between concentrations of SARS-CoV-2 RNA in wastewater and COVID-19 disease occurrence has used measurements from wastewater collected from an entire sewershed at wastewater treatment plants (Peccia et al., 2020; Graham et al., 2021; Prado et al., 2021; Wolfe et al., 2021a; Fernandez-Cassi et al., 2021), from smaller sub-sewersheds that encompass a cluster of buildings (Haak et al., 2022; Zambrana et al., 2022), or from buildings that serve as the primary location of residence such as dormitories (Betancourt et al., 2021; Langan et al., 2022), nursing homes (Davó et al., 2021; Schang et al., 2021), or prison facilities (Greenwald et al., 2021). However, wastewater monitoring for infectious disease targets at specific buildings such as K-12 schools, where individuals only spend a portion of their time, presents a unique set of challenges for application of wastewater-based epidemiology. This is because individuals may not use the toilets, showers, or sinks when they are in the buildings. At the same time, wastewater monitoring at these buildings may provide important information on disease occurrence that may help improve and guide local public health decisions.

During the 2021-2022 academic year, many K-12 schools employed optional regular clinical testing to identify individuals infected with SARS-CoV-2. One type of clinical surveillance scheme involved voluntary pooled clinical testing followed by individual testing. While participation in these programs can be high, it is not 100% (Faherty et al., 2021) and reporting delays associated with clinical test results can increase the time it takes to identify infected individuals. During this time, students may continue to be at high risk of exposure due to close proximity with other students in their daily activities. Wastewater, on the other hand, is a collective biological sample of the entire community using sinks, showers, and toilets on the premises. These samples can be collected and analyzed in less than 24 hours thereby overcoming limitations of clinical testing such as need for voluntary participation and reporting delays, while providing information on community infection rates to guide school policies or individuals’ choices on how to protect themselves.

Wastewater is a complex mixture containing both liquid and solid-phases. Previously, we showed that wastewater solids have a higher concentration of SARS-CoV-2 RNA than wastewater liquids on a mass equivalent basis in samples from wastewater treatment plants (Kim et al., 2022a). This is consistent with scientific literature that shows that viruses tend to partition to wastewater solids (Ye et al., 2016; Yin et al., 2018). At K-12 schools, this comparison has not been made to the best of our knowledge. Wastewater from clusters of buildings, like those at K-12 schools, spends a very short time in sewage lines between the source of wastewater such as toilets, sinks, or showers and the sampling location. We estimate less than 30 min based on preliminary use of fluorescence tracer at a sampling site compared to 2∼18 hours for wastewater to be transported to a nearby wastewater treatment plants (Wolfe et al., 2021b), and this short transit time might affect the partitioning behavior of viruses between the solid and liquid phase. Therefore, it is important to examine the relationship between the liquid and solid portions of wastewater in this setting to determine if solids are enriched with SARS-CoV-2 RNA even when wastewater is relatively fresh and represents an appropriate matrix for wastewater monitoring programs.

We conducted a review of the literature in August 2022 to identify peer-reviewed papers describing the use of wastewater for monitoring SARS-CoV-2 RNA at K-12 schools. The search was done using Google Scholar with key words “K-12 schools,” “wastewater,” and “SARS-CoV-2.” We identified two papers on this topic. Castro-Gutierrez et al. used the liquid portion of composite wastewater samples from sixteen K-12 schools and showed that they could detect SARS-CoV-2 RNA in school wastewater 47.3% of the time over the nine week period of their study; they did not compare their measurements with clinical data on disease occurrence (Castro-Gutierrez et al., 2022). Similarly, Crowe et al. collected grab samples of wastewater from schools and used the solid fraction of the collected wastewater to detect SARS-CoV-2 RNA over five weeks in three schools (Crowe et al., 2021); out of fourteen school-weeks when SARS-CoV-2 was detected by saliva testing, wastewater was positive for SARS-CoV-2 in twelve school-weeks.

In this study, we compare SARS-CoV-2 RNA concentrations recovered from solid and liquid fraction of wastewater samples to existing clinical pooled testing data from two K-12 schools in California. The goals of this work are to examine the relationship between SARS-CoV-2 RNA measurements in solid and liquid fraction of wastewater from a cluster of K-12 school buildings where individuals do not reside, and to verify the utility of the measurements in monitoring the school population for COVID-19 infections. We compared the viral RNA detection rate in wastewater to pooled clinical testing outcomes. The results of this work will inform the use of wastewater monitoring for SARS-CoV-2 at schools and other small settings.

## Materials and Methods

### Sample Collection

Wastewater samples were collected from two schools in California. Permission to sample was obtained from the superintendent of the school district. School A is a middle school with approximately 1000 individuals including 900 students. School B is a lower elementary school for students in Kindergarten to 2nd grade with approximately 400 individuals including 320 students. Both schools are located in the same county, within 5 km of each other. The two schools were chosen based on the availability of access points to sewage from the majority of the school. Wastewater was collected through access points that allowed us to sample before wastewater from the school buildings mixed with wastewater from other parts of the community (the access points were referred to as “sewage cleanout”); we estimate that wastewater spent < 30 min in the sewage pipes between where it entered the system and the sampling location. School A wastewater samples included wastewater from all buildings except for the gym and the music room, and School B wastewater samples included wastewater from the entire school.

Composite wastewater samples were collected using an autosampler (Teledyne ISCO, NE) programmed to collect 100 mL of wastewater every five minutes between 7:00 and 15:00 three times per week (Tuesdays, Wednesdays, and Fridays). This time period was selected to include students’ drop-off and pick-up times, which were around 8:00 and 14:00 respectively for both schools. Sampling was conducted for eight weeks from April 2022 to June 2022. Samples were retrieved by a field technician at the end of each day after samples were composited into a daily sample by the autosampler. The total volume for each sample varied according to the performance of the autosampler and ranged from 0 to 9.6 L. At the end of every sampling event, technicians decanted ∼8.5 L from the unmixed sample. The remaining 1 L of sample with a high solids content was immediately transported to the laboratory on ice. Once at the laboratory, the sample was stored at 4°C and analyzed within a week. Previous work indicated minimal degradation of viral RNA RT-PCR targets during this time frame (Roldan-Hernandez et al., 2022; Yang et al., 2022). The autosamplers malfunctioned twice and no sample was available at School A on April 19 and at School B on April 22. During the last week of this study, there was a national holiday on Monday; therefore samples were taken on Wednesday, Thursday, and Friday during this week.

### Pre-analytical processing

Wastewater samples were separated into a solid fraction and a liquid fraction by settling the sample in an Imhoff cone for one hour at 4°C (United States Environmental Protection Agency, 1999). The solid fraction was removed from the cone by decanting the liquid fraction into a sterile container and transferring the solid fraction to a different sterile container. Samples were processed using a modified version of Wolfe et al. (Wolfe et al., 2021b) The solid samples were dewatered by centrifuging at 24 000 xg for 30 minutes at 4°C and decanting the resulting supernatant. Solids included visible debris such as toilet paper. 0.225 g of solids were resuspended in DNA/RNA shield (Zymo Research, CA) so that the final concentration was 75 mg/mL, a concentration previously shown to minimize inhibition (Huisman et al., 2022). The DNA/RNA shield was spiked with bovine coronavirus (BCoV, Calf-guard Cattle Vaccine, PBS Animal Health, OH) at a concentration of 500 000 copies/mL, so that BCoV could be used as a spiked-in internal control. The resuspended sample was stored at 4°C overnight until nucleic acid extraction. If there were any leftover solids, dry weight of the dewatered solids was determined by drying the sample at 105°C for 24 hours.

Viruses from 45 mL of the previously separated liquid fraction were concentrated using electronegative filtration adapted from previous studies (LaTurner et al., 2021; Graham, Anderson & Boehm, 2021). The samples were centrifuged at 4 100 xg for 10 minutes at 4°C to eliminate any larger solids to prevent clogging. Sterile membranes (47 mm diameter; 0.45 µum pore size; Millipore HAWG047S6) were placed on sterile plastic filter funnels, wet with 1 mL of 1x phosphate buffered saline solution (PBS buffer, Gibco, NY) and the supernatant was transferred to the filter funnels. The sample was mixed with 1 mL of 1.25M MgCl_2_ and allowed to sit for five minutes. The sample was then vacuum filtered (approximate vacuum pressure of 17 kPa) through the sterile membrane. The vacuum was turned off to relieve the pressure and then 0.2 mL of DNA/RNA shield was placed onto the membrane, allowed to sit for five minutes, then aspirated from the filter using vacuum. The membrane was folded and placed in a bead beating tube included in the Qiagen AllPrep PowerViral DNA/RNA Kit (Qiagen, Germany) for extraction. 0.2 mL of DNA/RNA shield spiked with BCoV to a concentration of 500 000 copies/mL was added as a spiked-in internal control. Samples were stored at 4°C overnight until nucleic acid extraction.

### Nucleic Acid Extraction

For solid samples, 5/32” stainless steel grinder balls (OPS Diagnostics, NJ) were added to the suspension containing the solids and shaken with a bead beater (FastPrep-24 Tissue and Cell Homogenizer, MP Biomedicals, CA) to homogenize the sample. The homogenized solution was centrifuged at 5 250 xg for five minutes. Nucleic acids were extracted from the supernatant using the PowerViral Kit; manufacturer’s instructions were followed starting after the bead beating step. For liquid samples, membrane filters already added to the bead beating tubes in PowerViral Kit were processed as directed by the manufacturer. Qiacube (Qiagen, Germany) was used to extract nucleic acids for both solid and liquid samples using a custom program for the PowerViral Kit to elute 100 µL of RNA. Inhibitors were subsequently removed from the extracts using the Zymo OneStep-96 PCR Inhibitor Removal Kit (Zymo Research, Irvine, CA). Eluted RNA was used immediately after extraction to quantify RNA targets of interest, and remaining RNA was stored in -80°C. Extraction negative controls (DNA/RNA shield for solids and 1x PBS buffer for liquids) and positive controls (BCoV spiked in DNA/RNA shield) were processed using the same protocol every time a set of samples underwent nucleic acid extraction.

### Quantification

RNA targets were quantified using one-step droplet digital (dd)RT-PCR for two SARS-CoV-2 targets (N gene and S gene), BCoV, and pepper mild mottle virus (PMMoV), which acted as an endogenous internal recovery control and a fecal strength indicator (Wolfe et al., 2021a; Feng et al., 2021). A BioRad QX200 AutoDG droplet digital PCR system (BioRad, CA) was used along with the BioRad One-Step RT-ddPCR Advanced Kit for Probes (BioRad, CA). For SARS-CoV-2 targets, six replicate wells were used for each sample and merged in post-processing. For BCoV and PMMoV, two replicate wells were used for RNA templates diluted 1:10 in molecular grade water. Each plate included no-template PCR negative controls (water), extraction negative controls, and extraction positive controls (six replicate wells on SARS-CoV-2 plate and two on BCoV/PMMoV plate). Two positive PCR controls were run across all plates; the positive control consisted of RNA extracted from a nasopharynx swab of a high-titer COVID-19 patient from Stanford Hospital for SARS-CoV-2, direct extraction of BCoV vaccine diluted to ∼10^6^ cp/mL for BCoV, and synthetic DNA ultramer (Integrated DNA Technologies, Iowa) for PMMoV. Results were processed using QuantaSoft and QuantaSoft Analysis Pro (BoRad, CA) where replicate wells were merged and thresholded.

Each well needed to have at least 10,000 droplets generated to be included in the analysis. Three or more positive droplets across all replicate wells were required for a sample to be considered positive for the target. If there were less than 3 positive droplets, the sample was assigned as a non-detect (ND). Thresholds were chosen manually for each plate by setting a threshold for the no-template controls on the plate to have no more than 2 droplets above the threshold. Then the difference between this threshold and the average negative droplet fluorescence of the negative controls was set as the relative threshold difference for the plate. This difference was applied to all wells on the plate so that each well had a varying absolute threshold but a consistent relative threshold that reflected the fluctuation in the baseline negative droplet fluorescence.

The concentration per reaction was converted to copies per gram of dry weight or copies per mL of wastewater using dimensional analysis (formulas in Supplemental Article S1). Errors are standard deviations as “total error” from the instrument, which includes errors associated with variability among replicate wells and the Poisson distribution. If the sample did not have a dry weight measurement associated with it due to lack of adequate solid content in the sample, an average dry weight measurement from the school’s samples was used to calculate the corresponding copies per gram of dry weight at the end of the study period.

### Supplementary Wastewater Data

We obtained SARS-CoV-2 N gene and S gene RNA concentrations in addition to PMMoV RNA concentrations from settled solids at the wastewater treatment plant that processes the sewage from the sewershed these schools were part of. These wastewater treatment plant sample dates ranged from April 1, 2022 to June 10, 2022 to include the duration of this study. This data was acquired from the regional monitoring program to be used in a supplementary manner to examine association between school wastewater data and that of the surrounding community. These data have not been published previously but are available through the Stanford Digital Repository (https://purl.stanford.edu/km945rd8103). Methods used to acquire the regional monitoring data are described in detail by Wolfe et al. (Wolfe et al., 2021b) and briefly described in the Supplemental Article S1.

### Clinical Testing

Voluntary, weekly pooled clinical testing for COVID-19 was conducted at each school. Approximately 70% of the school attendees including students, teachers, and staff opted to participate in clinical testing. The schools were divided into cohorts where each cohort was tested on one day of the week (School A divided into three cohorts and School B into two).vNasal swabs of 7 to 17 individuals were collected and pooled together by the school and these samples were shipped overnight to be processed by a commercial lab using FDA-approved RT-PCR tests. The number of pools each day varied based on the number of individuals who sought testing but varied from 115 to 332 for School A and 66 to 486 for School B. Results of the pooled testing were communicated to the schools within 48 hours of specimen collection.

### Statistical analysis

Statistics were computed using RStudio (version 1.4.1106) and packages tidyverse, tidyr, zoo and stats. Nonparametric Mann-Whitney U test was used to test whether PMMoV RNA concentrations or PMMoV RNA concentration ratio in solid to liquid samples were significantly different between schools. Fisher’s exact test was used to compare detection frequency between solid and liquid samples. Bonferroni correction was not used in evaluating significance as all tests investigated a single hypothesis without multiple comparisons.

Nonparametric Kendall’s tau was used to assess association between RNA concentrations (PMMoV and SARS-CoV-2 N, S genes) in solid versus liquid samples. To account for technical variability of wastewater measurements, a bootstrapping approach was used where Kendall’s tau was calculated using 1000 resampling from a uniform distribution between upper and lower standard deviations of each sample concentration. Median tau and empirical p-values were calculated from this resampling (Wolfe et al., 2021a). For samples that were reported as non-detects (NDs), bootstrapping bounds were defined as zero for the lower standard deviation and the lowest theoretical measurement limit for the upper standard deviation. The lowest theoretical measurement limit for each sample was calculated by determining the concentration of each sample if only three droplets across all replicate wells were positive (our criteria to be considered to have detectable concentrations of the target). Because every sample has a different number of generated droplets and dry weight associated with the sample, the lowest theoretical measurement limit varies from sample to sample (values provided in results section).

Empirical relationships between RNA concentrations of matched solid and liquid samples were established with linear regression of log-transformed data to derive slopes and y-intercepts. Only samples that were positive for the SARS-CoV-2 target in both matched solid and liquid samples were used to investigate this relationship.

To compare the detection results of wastewater sampling and pooled clinical testing, we compared complete sets of weekly pooled clinical test results to wastewater samples collected the day of or one day after specimen collection as shown in Fig. 1. This matching scheme for wastewater and clinical testing data was chosen to account for the time it takes to run clinical tests and identify an individual infected with COVID-19 (up to 48 hours) to be taken out of the school system. We assumed that until identified, the infected individual was still contributing to the school wastewater. Weekly sets of pooled tests were considered as a unit of analysis as these represent clinical testing results for the entire school, and the entire school is the population theoretically represented by the wastewater samples. Detection of SARS-CoV-2 RNA in Tuesday or Wednesday wastewater samples was compared to sets of three pooled testing results at School A, done in mutually exclusive cohorts on Monday, Tuesday, and Wednesday of each week. Similarly, detection of SARS-CoV-2 RNA in Tuesday or Wednesday wastewater samples was compared to sets of two pooled testing results at School B, done in mutually exclusive cohorts on Monday and Tuesday of each week. Fisher’s exact test was used to compare solid and liquid in their ability to accurately reflect results of the positive pooled testing.

**Figure 1.**
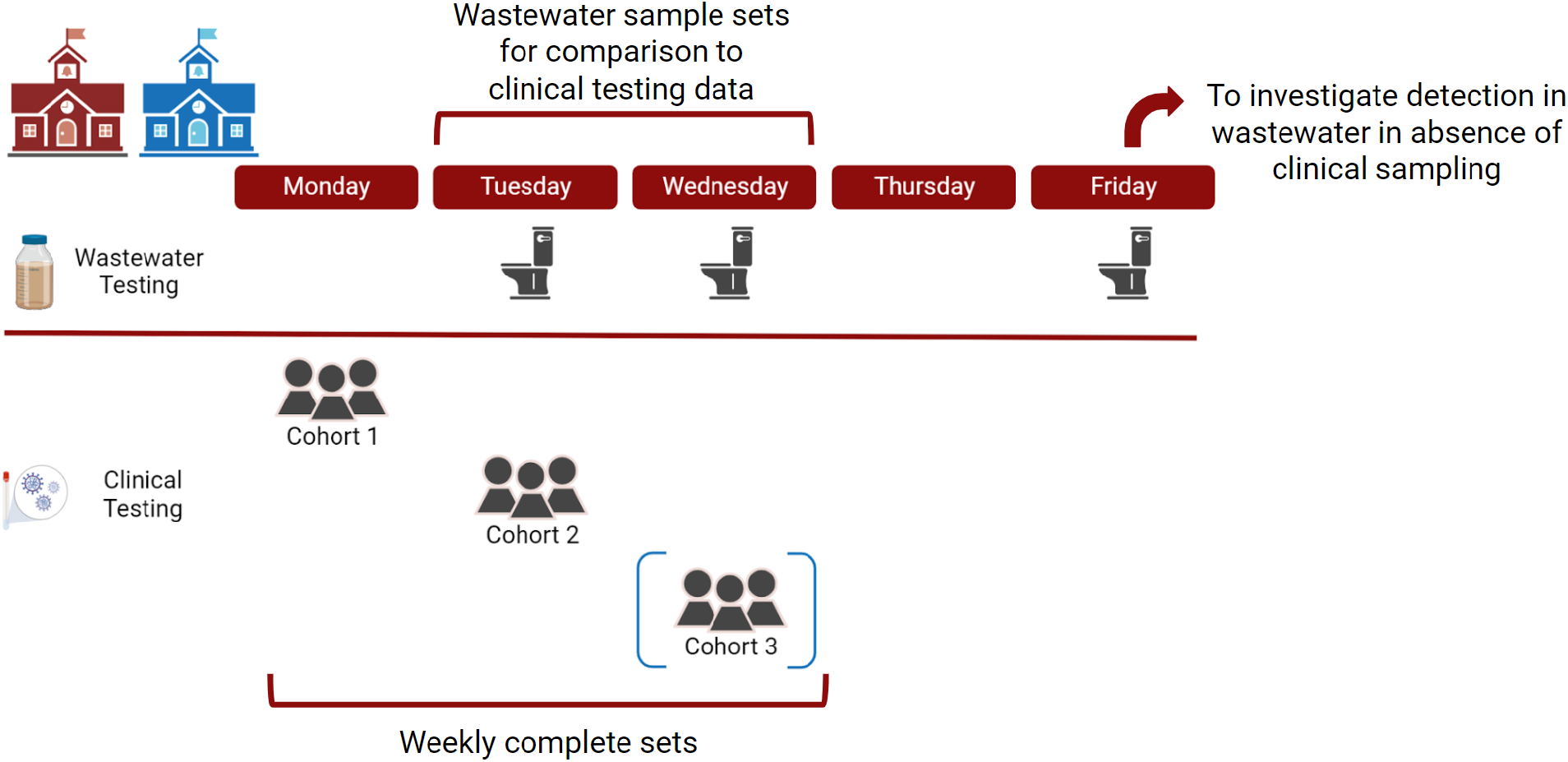
Sampling scheme for surveillance at two schools in the study. For clinical pooled testing, School A had three mutually exclusive cohorts and School B had two mutually exclusive cohorts, indicated by the blue brackets around cohort 3.

## Results

### Quality assurance (QA)/quality control (QC)

Negative and positive extraction and PCR controls were ND and positive, respectively. Recovery rates of BCoV indicated no RNA extraction failures (recovery rate > 10%). One sample in School A did not have a detectable concentration of PMMoV; we considered this sample to have experienced an RNA extraction failure and excluded it from further analysis. After removal of that sample, there were 91 samples (45 solid and 46 liquid samples) from the two schools remaining for inclusion in our analyses.

### Measurement overview

Across solid samples, PMMoV ranged from 2.6 × 10^6^ to 1.9 × 10^9^ copies g^-1^ dry weight (hereafter referred to as cp g^-1^, median = 7.8 × 10^7^); across liquid samples, PMMoV ranged from 5.4 × 10^2^ to 9.8 × 10^5^ copies mL^-1^ wastewater (hereafter referred to as cp mL^-1^, median = 1.4 × 10^4^) (Fig. S1). PMMoV concentrations were not different between schools for solids or liquids (two Mann-Whitney U tests, p-value for solids = 0.10, for liquid = 0.73). Median ratio of PMMoV concentrations in matched solid to liquid samples across both schools was 4.1 × 10^4^ mL g^-1^ (n = 45, range 3.8 × 10^2^ to 8.2 × 10^4^ mL g^-1^). Ratios were different between schools with School B having a higher median ratio (4.8 × 10^4^ mL g^-1^) compared to School A (3.4 × 10^3^ mL g^-1^) (Mann-Whitney U test, p-value = 0.03). PMMoV concentrations in matched solid and liquid samples were positively and significantly correlated at the two schools as both aggregated data and at individual school level (Table 1).

**Table 1.**
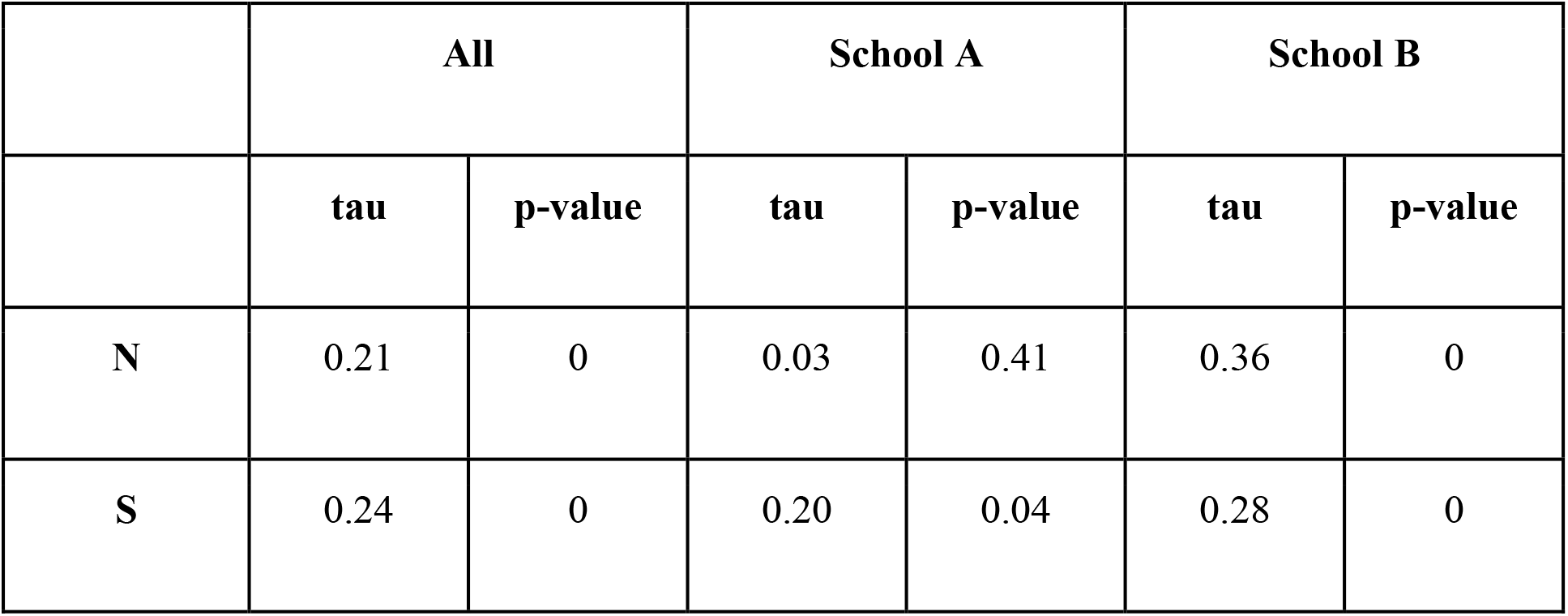
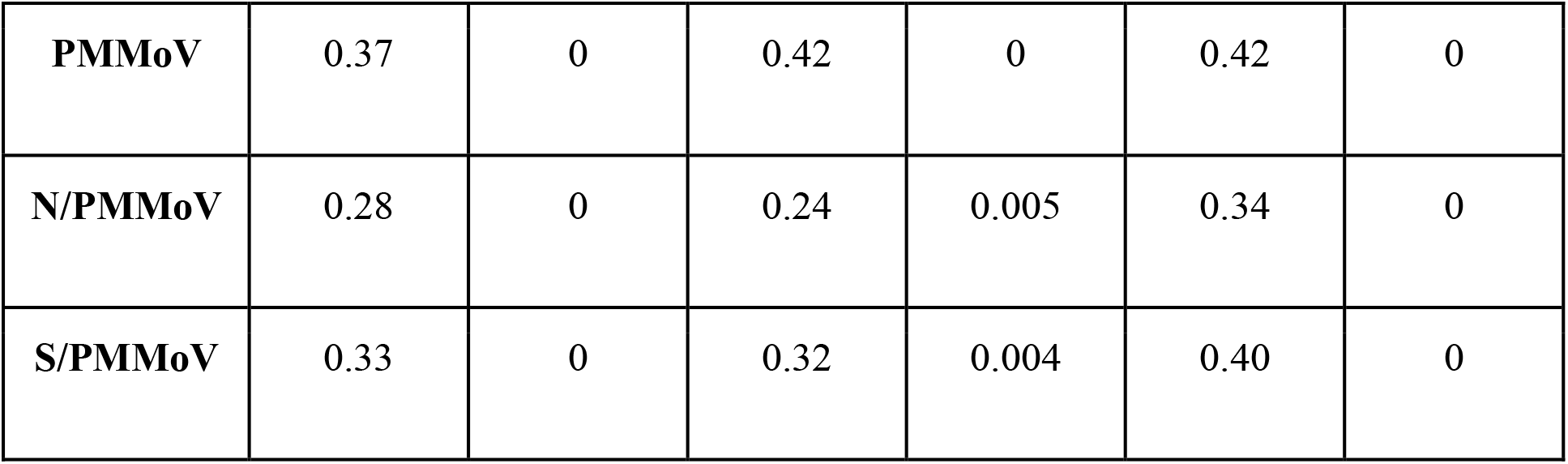
Median Kendall’s tau between wastewater SARS-CoV-2 RNA in matched solid and liquid samples. 1000 instances of Kendall’s tau were calculated by sampling between upper and lower confidence intervals of each measured concentration of SARS-CoV-2 RNA. Empirical p-value was calculated as a percentage of Kendall’s tau that resulted in a negative value.

In the solid and liquid fractions of each sample, N and S gene targets of SARS-CoV-2 were measured (Fig. 2). SARS-CoV-2 RNA gene concentrations in solids ranged from ND to 4.4 × 10^4^ cp g^-1^ (N) and from ND to 1.1 × 10^5^ cp g^-1^ (S). Across liquid samples, SARS-CoV-2 RNA concentrations ranged from ND to 41 cp mL^-1^ (N) and from ND to 6 cp mL^-1^ (S) (Fig. S2). The lowest theoretical measurement limit calculated for each sample ranged from 3.3 × 10^3^ to 1.7 × 10^4^ cp g^-1^ (median = 9.7 × 10^3^) for solid samples and 0.2 cp mL^-1^ to 0.5 cp mL^-1^ (median = 0.3) for liquid samples (Fig. S3). Lowest observed measurements for solid samples were 8.1 × 10^3^ (N) and 8.9 × 10^3^ (S) cp g^-1^; for liquid samples were 0.3 cp mL^-1^ (N and S). When compared to the larger sewershed that the schools were part of, the SARS-CoV-2 concentrations at each of the schools, only School B had a positive and significant association (Fig. S4, Table S1). All wastewater data presented in this paper can be found in the Stanford Digital Repository (https://purl.stanford.edu/sy647tw8455).

**Figure 2.**
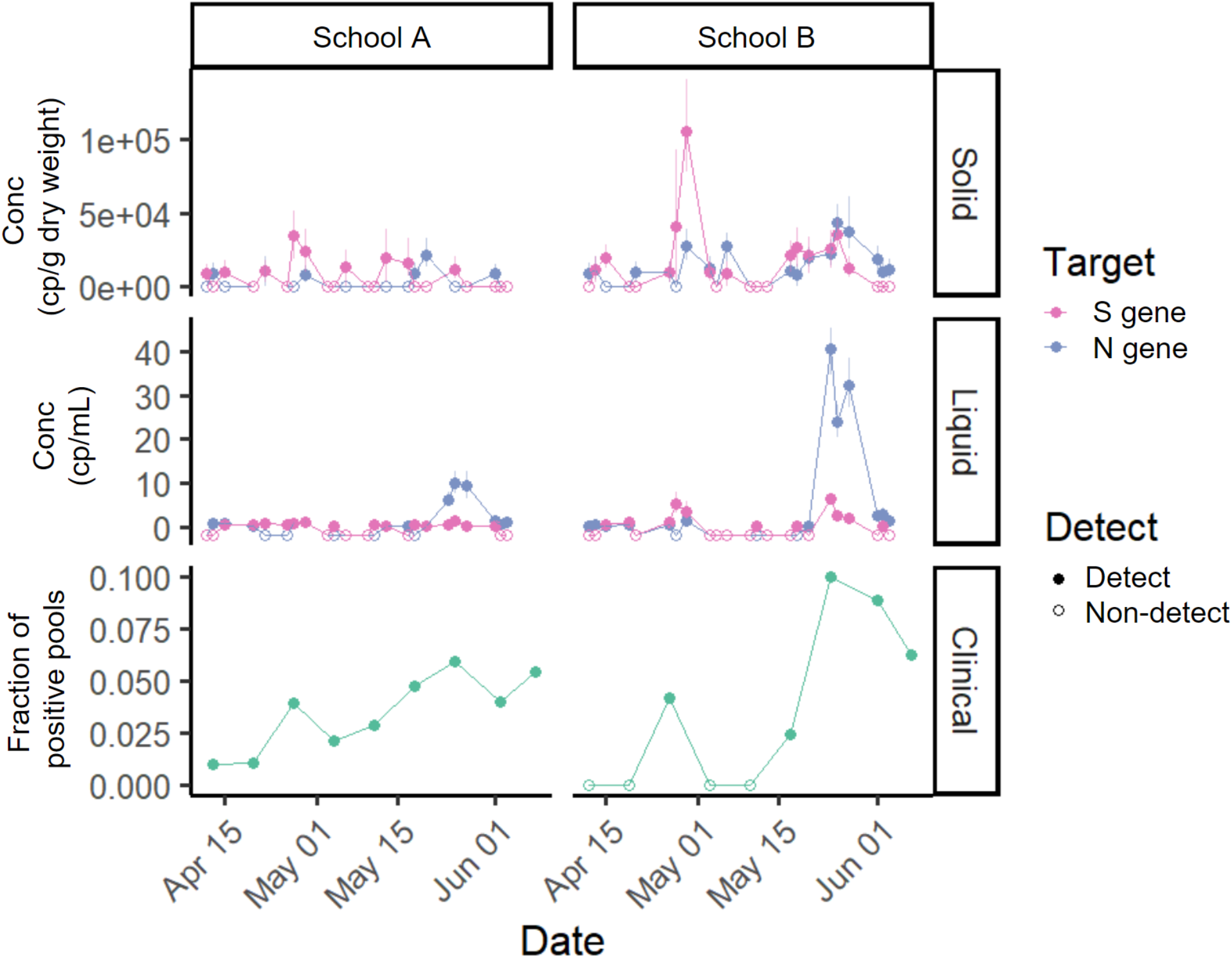
Time series of measured SARS-CoV-2 in the study. SARS-CoV-2 targets N or S measured in solid samples in cp g^-1^ dry weight (top), concentration measured in liquid samples in cp mL^-1^ (middle), and fraction of positive pools for each of the schools (bottom) in the study over eight weeks. Each wastewater data point represents SARS-CoV-2 RNA concentration for a single sample. Samples below the lower measurement limit are shown as empty circles just below 0 to aid with visualization. For clinical samples, empty circles represent no positive pools. The same time series showing wastewater measurements of N and S normalized by PMMoV can be found in the SI (Fig. S7).

N and S were significantly correlated within both solid (R^2^ = 0.12, slope = 0.22, p-value = 0.01) and liquid measurements (R^2^ = 0.39, slope = 3.97, p-value = 2.2×10-6) (Fig. S5). Although they were positively associated, N and S measurements were not always in agreement; detection of N and S gene were in agreement (i.e. both detected or both undetected) in 62% of samples and they were in disagreement (i.e. one gene detected but the other gene undetected) in 38% samples across both solid and liquid samples. When separated by matrix, N and S gene detection agreement remained similar (63% in liquid and 60% in solid). In subsequent analyses that involved examining the relationship between SARS-CoV-2 RNA concentration in solid and liquid samples, the two targets were analyzed independently. When comparing the detection rate of wastewater samples and clinical pooled testing, an individual solid or liquid wastewater sample was considered positive for SARS-CoV-2 if either one of the targets (S or N) was detected.

### Relationship between SARS-CoV-2 RNA in solids and liquids

Out of the 45 matched liquid and solid samples, 31 solid samples were positive and 14 samples were negative for SARS-CoV-2 RNA; 34 liquid samples were positive and 11 samples were negative for SARS-CoV-2 RNA (Table 2). There is no significant difference between detection frequency in solid versus liquid samples (Fisher’s exact test statistic value of 0.64). The result did not change when samples from each school were examined separately. Solid and liquid samples agreed on detection in the majority of samples (67%) but there were samples that were positive for SARS-CoV-2 RNA in the solids but negative in the liquid and vice versa. School B samples had a higher rate of agreement (74%) between solids and liquids, but this rate was not statistically different from the rate for School A samples (59%) (Fisher’s exact test statistic value of 0.35).

**Table 2.**
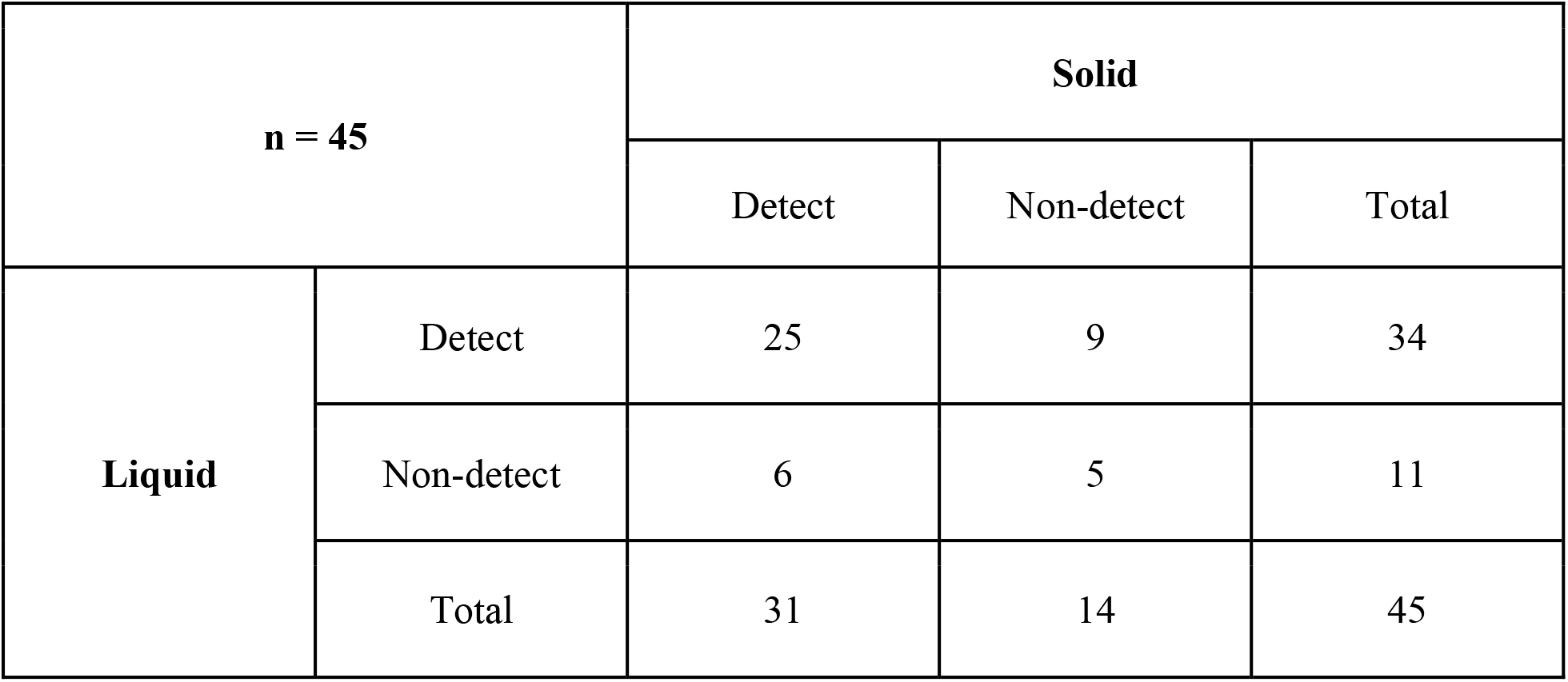
Detection frequency of solid and liquid samples aggregated across schools and the entire study period.

| **n = 45** |  | **Solid** |  |  |
| --- | --- | --- | --- | --- |
|  |  | Detect | Non-detect | Total |
| **Liquid** | Detect | 25 | 9 | 34 |
|  | Non-detect | 6 | 5 | 11 |
|  | Total | 31 | 14 | 45 |

The median ratio of SARS-CoV-2 RNA concentrations in matched solid and liquid samples where both matrices were positive for the SARS-CoV-2 N gene was 8.6 × 10^3^ mL g^-1^ (n = 15, min 5.6 × 10^2^ mL g^-1^, max 6.5 × 10^4^ mL g^-1^) and for the S gene was 1.6 × 10^4^ mL g^-1^ (n = 14, min 4.0 × 10^3^ mL g^-1^, max 9.3 × 10^4^ mL g^-1^) (Table 3). The ratios of N gene and S gene are not statistically different (Mann Whitney U test, p-value = 0.16). SARS-CoV-2 RNA concentrations in matched solid and liquid samples were positively and significantly correlated for both N (median Kendall’s tau = 0.21, empirical p-value = 0) and S gene (median Kendall’s tau = 0.24, empirical p-value = 0) as aggregated data. At the individual school level, SARS-CoV-2 concentrations in matched solid and liquid samples for each school were positively and significantly correlated (median Kendall’s tau > 0, empirical p-value < 0.05) with the exception of N gene for School A (Kendall’s tau = 0.03, empirical p-value = 0.41). When SARS-CoV-2 RNA concentration was normalized by PMMoV, SARS-CoV-2 concentrations in matched solid and liquid samples for each school were positively and significantly correlated for both targets (median Kendall’s tau > 0, empirical p-value < 0.05) (Table 1).

**Table 3.** Ratio of RNA in wastewater in matched solid and liquid samples aggregated across schools and the entire study period. Number of matched samples (n),and 25th percentile, median, and 75th percentile values for each target gene and their normalized values are reported. Only samples that had a positive detection in both solid and liquid samples were included in the calculation of the ratio.

|  | **n** | **25th Percentile** | **Median** | **75th Percentile** |
| --- | --- | --- | --- | --- |
| **N gene** | 15 | $4.9 \times 10^3$ | $8.6 \times 10^3$ | $2.3 \times 10^4$ |
| **S gene** | 14 | $8.7 \times 10^3$ | $1.6 \times 10^4$ | $3.6 \times 10^4$ |
| **PMMoV** | 45 | $2.8 \times 10^4$ | $4.1 \times 10^4$ | $1.1 \times 10^4$ |
| **N/PMMoV** | 15 | 0.53 | 1.8 | 3.4 |
| **S/PMMoV** | 14 | 1.9 | 3.6 | 4.7 |

To derive an empirical relationship between the log10-transformed solids and liquid concentrations, we used linear regression where y is the log10-transformed solid concentration (cp g^-1^) and x is the log10-transformed liquid concentration (cp mL^-1^), as in a Freundlich isotherm model (Schwarzenbach, Gschwend & Imboden, 2017): 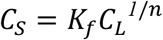 where *C*_*S*_ is RNA concentration in the solid samples, *C*_*L*_ is RNA concentration in liquid samples, *K*_*f*_ is the Freuidlich’s constant, and *1/n* is the exponent of non-linearity. For this analysis, we used only data points that had SARS-CoV-2 RNA concentration detected in both solid and liquid samples instead of substituting ND with other values. This was done to prevent the substituted values from having an impact on the derived relationship. Data from the two schools were examined in aggregate. For the N gene, the slope was 0.21 and the y-intercept was 4.1, indicating n = 4.8 and K_f_ = 10^4^ mL g^-1^ in the Freundlich model. For the S gene, the slope was 0.29 and the y-intercept was 4.3, indicating n = 3.4 and K_f_ = 10^4^ mL g^-1^ (Fig. 3, Table 4).

**Table 4.** Linear regression coefficients between log_10_-transformed SARS-CoV-2 concentration in matched solid and liquid samples. The slope and intercept are from a linear regression of Y = slope * X + intercept where X = log_10_- transformed liquid concentration and Y = log_10_-transformed solid concentration. Number of samples used in the regression is shown in n. Adj R^2^ (square of the Pearson correlation coefficient adjusted for degrees of freedom) and the p-value (showing statistical significance of the coefficients) are outputs from the lm function in R.

| **Target** | **n** | **Slope** | **Intercept** | **Adj R<sup>2</sup>** | **p-value** |
| --- | --- | --- | --- | --- | --- |
| **N** | 15 | 0.21 | 4.1 | 0.34 | 0.01 |
| **S** | 14 | 0.29 | 4.3 | 0.11 | 0.13 |
| **PMMoV** | 45 | 0.75 | 4.7 | 0.40 | < 0.01 |

**Figure 3.**
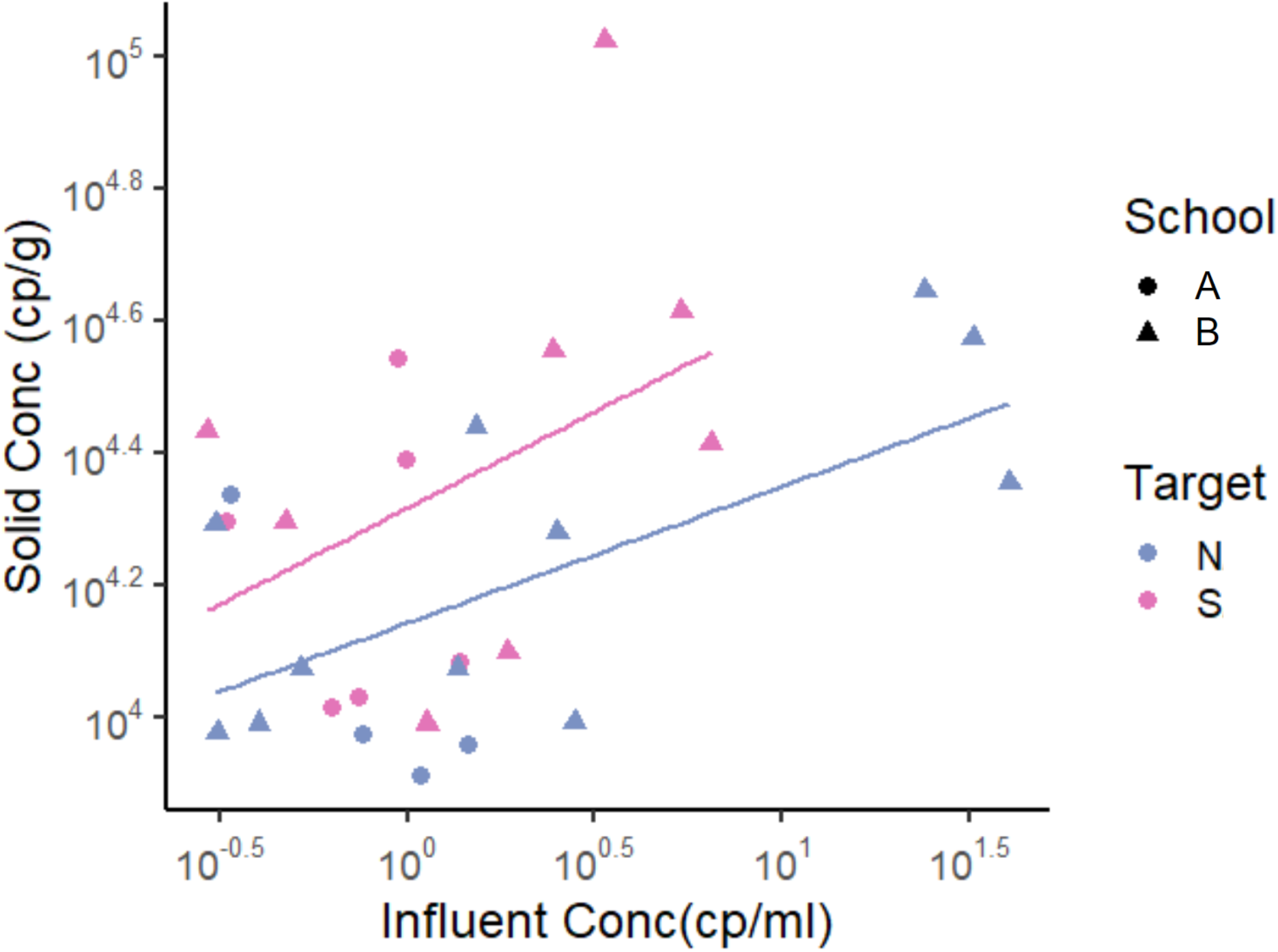
SARS-CoV-2 RNA concentrations in matched solid and liquid samples. Only data points that had SARS-CoV-2 RNA concentration detected in both solid and liquid samples were used. Each data point represents SARS-CoV-2 RNA concentration from a single sample. Note that data are displayed in log10-scale format for ease of visualization. Pairwise linear regression lines are shown for each target in their respective color.

### Comparison with pooled testing

Out of 16 school-weeks total (eight weeks at two schools) where pooled testing was completed, COVID-19 infection was confirmed in pooled testing for 12 of the school-weeks. During these 12 school-weeks, 9 sets of the solid samples (75%) and 12 sets of the liquid samples (100%) included positive detections of SARS-CoV-2 RNA in wastewater. The difference between the detection rate of solid (75%) and liquid (100%) samples to detect SARS-CoV-2 RNA in wastewater when pooled testing was positive is not significantly different (Fisher’s exact test, p-value = 0.22). In the 4 school-weeks of negative pooled clinical specimen testing, 3 weeks of solid samples (75%) and 3 sets of liquid samples (75%) included positive detection of SARS-CoV-2 RNA in wastewater (Fig. 4).

**Figure 4.**
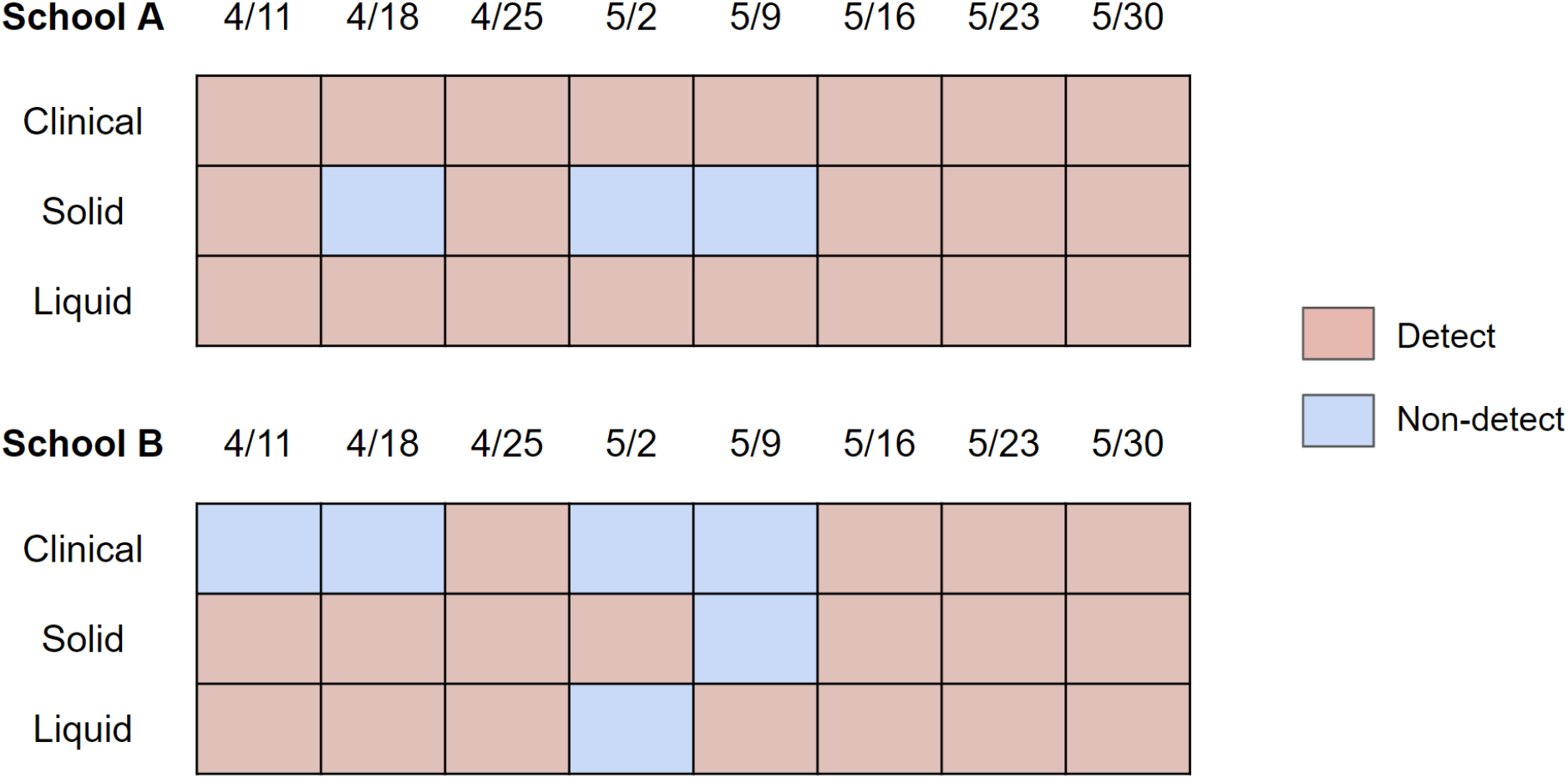
Detection/non-detection compared between solid and liquid fraction of wastewater, and clinical pooled testing results for days that pooled testing was conducted. The dates correspond to the first day of the week for the collected sets of samples. Clinical samples are sets of three (for School A) or two (for School B) cohorts of weekly pooled testing. Solid and liquid samples are sets of Tuesday and Wednesday samples. The result was shown as “Detect” if either of Tuesday or Wednesday samples were positive for SARS-CoV-2. A more specific breakdown of detection rates is shown in Fig. S6.

On Fridays, clinical specimens were not collected but wastewater samples were. Out of 15 school-days total (eight Fridays at two schools with one failed sampling event), 12 solid samples (80%) and 12 liquid samples (80%) were positive for SARS-CoV-2 RNA (Fig. S6), indicating the possibility that the week’s clinical sampling may not have removed all of the individuals affected with COVID-19 from the campus.

## Discussion

In this study we compared detection of SARS-CoV-2 RNA in wastewater to detection of the virus in pooled clinical specimens. When pooled clinical specimens were positive, indicating that a specimen from at least one individual in the pool was infected with SARS-CoV-2, most wastewater samples collected within two days of specimen collection were positive for SARS-CoV-2 RNA. This indicates that wastewater samples, both solid and liquid fraction, can be used to identify the presence of SARS-CoV-2 infection in a school community.

A large fraction (83% for both solid and liquid samples) of wastewater samples were positive for SARS-CoV-2 RNA on days when pooled clinical specimens were negative or in absence of clinical testing. While this could possibly be from residual shedding in stools of individuals that have recovered from COVID-19 (Natarajan et al., 2022), it may also indicate the presence of infected individuals in the school. A voluntary clinical testing program for which not all individuals are required to provide specimens cannot detect all infected individuals, and may yield false negative results. On the other hand, one wastewater sample provides an indication of COVID-19 infections representative of all who use the sewer system on the school campus. Wastewater can also be easily accessed and sampled frequently to provide insight into infection dynamics without asking the community to engage in behavior changes (i.e., testing). This highlights the potential for wastewater monitoring to supplement more traditional methods of measuring COVID-19 occurrence in small community settings like K-12 schools.

We compared SARS-CoV-2 RNA concentration in solid and liquid fractions of wastewater at buildings that do not serve as the main location of residence for its community members.

Compared to the results from our previous study where we investigated wastewater collected from an entire sewershed at wastewater treatment plants (Kim et al., 2022a), the overall RNA concentration of both PMMoV and SARS-CoV-2 were lower with a larger variation at the two K-12 schools. This is to be expected from a smaller sewershed with smaller population size (Gibas et al., 2021). However, other general trends were consistent with observations from wastewater treatment plants in that 1) solid and liquid fraction showed comparable detection frequency for SARS-CoV-2 RNA, and 2) there was approximately a 10^3^-10^4^ higher concentration in the solid fraction of both SARS-CoV-2 RNA and PMMoV RNA on an equivalent mass basis. This indicates that SARS-CoV-2 RNA is preferentially associated with the solid fraction of wastewater at the K-12 setting.

One limitation of this study in comparing pooled clinical testing to wastewater sampling is that we were unable to collect a Monday wastewater sample to match clinical specimen collection completed on Monday due to operational challenges in the field. However, we did have an approximate clinical testing schedule for each of the schools, which showed that follow-up individual testing took place when pooled testing results were announced, approximately 48 to 72 hours after pooled testing took place. This means that individuals that participated in the Monday pooled testing would still most likely be in school on Tuesday when we were able to collect a wastewater sample. There were limitations associated with the pooled clinical testing data itself. Participation of the pooled testing was voluntary, and therefore it is possible to have false negative results for the school even though the majority of the school opted to participate. In addition, the pooled clinical testing data was used only qualitatively (i.e. detect or non-detect) as a single positive pool may have multiple infected individuals, especially considering that pools were created based on location proximity (i.e. students in the same classroom are put together in one pool).

There were a few challenges associated with working in a K-12 sewershed that should be considered. Rieckermann et al. analyzed 60 tracer experiments in 37 different sewers to show that dispersion in sewers is generally very small with little variation (Rieckermann et al., 2005). This implies that sewer inputs containing the viral RNA will act as discrete pulses passing through the sampling point and that this signal could be missed without frequent sampling. The closer the sampling is to the toilets, sinks, and showers that connect to the sewer, the lower the probability of collecting a representative sample without a comprehensive sampling scheme (Wade et al., 2022) as we had. Second, schools actively seek to remove individuals infected with COVID-19, which leads to low concentrations of SARS-CoV-2 RNA in wastewater, as shown in this study. Therefore sensitive methods, including those focused on the solids component of wastewater which is enriched in viral RNA, could be beneficial to detect the viral RNA in wastewater. Lastly, if the infected individual does not use a toilet during their time at the school, they will not contribute to the wastewater leading to a potentially false negative for wastewater. There are other ways for a biological sample (i.e. saliva, respiratory fluids) to enter the sewer to contribute to the observed SARS-CoV-2 RNA concentration in wastewater. However, Crank et al. showed that even at building level, stool was the most probable primary contributor to the SARS-CoV-2 RNA concentrations in wastewater (Crank et al., 2022).

Based on these challenges, we provide the following recommendations for other researchers and school administrators interested in wastewater monitoring applications for COVID-19 surveillance. First, to account for how close the wastewater sampling point is to the source, a very frequent compositing scheme is needed. Autosamplers with short intervals between its sampling events (as in this study) or continuous samplers would be suitable. Second, to effectively detect SARS-CoV-2 RNA at low concentrations, multiple PCR targets based on different regions of the SARS-CoV-2 genome should be used. Ahmed et al. showed that combining multiple assays can increase SARS-CoV-2 RNA detection sensitivity (Ahmed et al., 2022). As recommended by Ahmed et al. (Ahmed et al., 2022), we took the presence of at least one of the targets to indicate presence of SARS-CoV-2 RNA. While the detection rate, as a percentage, did not differ greatly when only one gene was considered, almost ⅔ of the samples that tested positive for SARS-CoV-2 RNA showed detection of one gene and not the other. Additionally, if ddPCR is used, multiple replicate PCR wells should be used and merged to increase the reaction volume associated with the sample and in turn increase the chance of detecting SARS-CoV-2 RNA (Kim et al., 2022b). Lastly, homogenizing solids obtained from wastewater is required. In our solid samples, there were large amounts of debris such as toilet paper that contributed to the heterogeneity of the solid samples. Either a sieve in front of the sampling port that could prevent such debris from entering the sample collection chamber or an additional step during pre-analytical steps to homogenize the sample may improve consistency in SARS-CoV-2 RNA detection in solids.

## Conclusions

SARS-CoV-2 RNA in wastewater measured in both solid and liquid fraction of wastewater at K-12 schools were able to detect the virus with comparable detection frequency. Wastewater solid fraction had a higher concentration than the liquid fraction in equivalent mass, consistent with previous studies conducted at larger wastewater treatment plants. Most wastewater samples were positive on days when pooled clinical sampling was positive; there were also positive wastewater samples when pooled testing was negative or in absence of clinical testing, suggesting the presence of individuals on campus shedding viral RNA who did not participate in testing, or convalescing individuals still shedding viral RNA. We provide recommendations for working in a sewershed serving a small population with low concentrations of the virus in wastewater. In this work, the two schools were chosen with care after verifying that there was a convenient access point and only one outlet for sewage. Further work should be done to determine how to integrate wastewater surveillance to schools with a more complex sewer system and multiple outlets.

## Data Availability

All data produced are available online at the Stanford Digital Repository (links in the body of the main article).

https://purl.stanford.edu/km945rd8103

https://purl.stanford.edu/sy647tw8455

## Acknowledgements

We thank the anonymous school district for their help with sampling and support of the study and those involved in the sampling and transportation of the samples. We also thank Concentric by Ginkgo for program management of the pooled testing and wastewater testing, donation of reagents, and additional assistance they provided throughout the study.

## References

Ahmed W, Bivins A, Metcalfe S, Smith WJM, Ziels R, Korajkic A, McMinn B, Graber TE, Simpson SL. 2022. RT-qPCR and ATOPlex sequencing for the sensitive detection of SARS-CoV-2 RNA for wastewater surveillance. Water Research 220:118621. DOI: 10.1016/j.watres.2022.118621.

Betancourt WQ, Schmitz BW, Innes GK, Prasek SM, Pogreba Brown KM, Stark ER, Foster AR, Sprissler RS, Harris DT, Sherchan SP, Gerba CP, Pepper IL. 2021. COVID-19 containment on a college campus via wastewater-based epidemiology, targeted clinical testing and an intervention. Science of The Total Environment 779:146408. DOI: 10.1016/j.scitotenv.2021.146408.

Castro-Gutierrez V, Hassard F, Vu M, Leitao R, Burczynska B, Wildeboer D, Stanton I, Rahimzadeh S, Baio G, Garelick H, Hofman J, Kasprzyk-Hordern B, Kwiatkowska R, Majeed A, Priest S, Grimsley J, Lundy L, Singer AC, Di Cesare M. 2022. Monitoring occurrence of SARS-CoV-2 in school populations: A wastewater-based approach. PLOS ONE 17:e0270168. DOI: 10.1371/journal.pone.0270168.

Crank K, Chen W, Bivins A, Lowry S, Bibby K. 2022. Contribution of SARS-CoV-2 RNA shedding routes to RNA loads in wastewater. Science of The Total Environment 806:150376. DOI: 10.1016/j.scitotenv.2021.150376.

Crowe J, Schnaubelt AT, SchmidtBonne S, Angell K, Bai J, Eske T, Nicklin M, Pratt C, White B, Crotts-Hannibal B, Staffend N, Herrera V, Cobb J, Conner J, Carstens J, Tempero J, Bouda L, Ray M, Lawler JV, Campbell WS, Lowe J-M, Santarpia J, Bartelt-Hunt S, Wiley M, Brett-Major D, Logan C, Broadhurst MJ. 2021. Assessment of a Program for SARS-CoV-2 Screening and Environmental Monitoring in an Urban Public School District. JAMA Network Open 4:e2126447. DOI: 10.1001/jamanetworkopen.2021.26447.

Davó L, Seguí R, Botija P, Beltrán MJ, Albert E, Torres I, López-Fernández PÁ, Ortí R, Maestre JF, Sánchez G, Navarro D. 2021. Early detection of SARS-CoV-2 infection cases or outbreaks at nursing homes by targeted wastewater tracking. Clinical Microbiology and Infection 27:1061–1063. DOI: 10.1016/j.cmi.2021.02.003.

Faherty LJ, Master BK, Steiner ED, Kaufman JH, Predmore Z, Stelitano L, Leschitz JT, Phillips B, Schwartz HL, Wolfe R. 2021. COVID-19 Testing in K-12 Schools.

Feng S, Roguet A, McClary-Gutierrez JS, Newton RJ, Kloczko N, Meiman JG, McLellan SL. 2021. Evaluation of Sampling, Analysis, and Normalization Methods for SARS-CoV-2 Concentrations in Wastewater to Assess COVID-19 Burdens in Wisconsin Communities. ACS ES&T Water 1:1955–1965. DOI: 10.1021/acsestwater.1c00160.

Fernandez-Cassi X, Scheidegger A, Bänziger C, Cariti F, Tuñas Corzon A, Ganesanandamoorthy P, Lemaitre JC, Ort C, Julian TR, Kohn T. 2021. Wastewater monitoring outperforms case numbers as a tool to track COVID-19 incidence dynamics when test positivity rates are high. Water Research 200:117252. DOI: 10.1016/j.watres.2021.117252.

Gibas C, Lambirth K, Mittal N, Juel MAI, Barua VB, Roppolo Brazell L, Hinton K, Lontai J, Stark N, Young I, Quach C, Russ M, Kauer J, Nicolosi B, Chen D, Akella S, Tang W, Schlueter J, Munir M. 2021. Implementing building-level SARS-CoV-2 wastewater surveillance on a university campus. Science of The Total Environment 782:146749. DOI: 10.1016/j.scitotenv.2021.146749.

Graham KE, Anderson CE, Boehm AB. 2021. Viral pathogens in urban stormwater runoff: Occurrence and removal via vegetated biochar-amended biofilters. Water Research 207:117829. DOI: 10.1016/j.watres.2021.117829.

Graham KE, Loeb SK, Wolfe MK, Catoe D, Sinnott-Armstrong N, Kim S, Yamahara KM, Sassoubre LM, Mendoza Grijalva LM, Roldan-Hernandez L, Langenfeld K, Wigginton KR, Boehm AB. 2021. SARS-CoV-2 RNA in Wastewater Settled Solids Is Associated with COVID-19 Cases in a Large Urban Sewershed. Environmental Science & Technology 55:488–498. DOI: 10.1021/acs.est.0c06191.

Greenwald HD, Kennedy LC, Hinkle A, Whitney ON, Fan VB, Crits-Christoph A, Harris-Lovett S, Flamholz AI, Al-Shayeb B, Liao LD, Beyers M, Brown D, Chakrabarti AR, Dow J, Frost D, Koekemoer M, Lynch C, Sarkar P, White E, Kantor R, Nelson KL. 2021. Tools for interpretation of wastewater SARS-CoV-2 temporal and spatial trends demonstrated with data collected in the San Francisco Bay Area. Water Research X 12:100111. DOI: 10.1016/j.wroa.2021.100111.

Haak L, Delic B, Li L, Guarin T, Mazurowski L, Dastjerdi NG, Dewan A, Pagilla K. 2022. Spatial and temporal variability and data bias in wastewater surveillance of SARS-CoV-2 in a sewer system. Science of The Total Environment 805:150390. DOI: 10.1016/j.scitotenv.2021.150390.

Huisman JS, Scire J, Caduff L, Fernandez-Cassi X, Ganesanandamoorthy P, Kull A, Scheidegger A, Stachler E, Boehm AB, Hughes B, Knudson A, Topol A, Wigginton KR, Wolfe MK, Kohn T, Ort C, Stadler T, Julian TR. 2022. Wastewater-Based Estimation of the Effective Reproductive Number of SARS-CoV-2. Environmental Health Perspectives 130:057011. DOI: 10.1289/EHP10050.

Kim S, Kennedy LC, Wolfe MK, Criddle CS, Duong DH, Topol A, White BJ, Kantor RS, Nelson KL, Steele JA, Langlois K, Griffith JF, Zimmer-Faust AG, McLellan SL, Schussman MK, Ammerman M, Wigginton KR, Bakker KM, Boehm AB. 2022a. SARS-CoV-2 RNA is enriched by orders of magnitude in primary settled solids relative to liquid wastewater at publicly owned treatment works. Environmental Science: Water Research & Technology:10.1039.D1EW00826A. DOI: 10.1039/D1EW00826A.

Kim S, Wolfe MK, Criddle CS, Duong DH, Chan-Herur V, White BJ, Boehm AB. 2022b. Effect of SARS-CoV-2 digital droplet RT-PCR assay sensitivity on COVID-19 wastewater based epidemiology. Infectious Diseases (except HIV/AIDS). DOI: 10.1101/2022.04.17.22273949.

Langan LM, O’Brien M, Rundell ZC, Back JA, Ryan BJ, Chambliss CK, Norman RS, Brooks BW. 2022. Comparative Analysis of RNA-Extraction Approaches and Associated Influences on RT-qPCR of the SARS-CoV-2 RNA in a University Residence Hall and Quarantine Location. ACS ES&T Water:acsestwater.1c00476. DOI: 10.1021/acsestwater.1c00476.

LaTurner ZW, Zong DM, Kalvapalle P, Gamas KR, Terwilliger A, Crosby T, Ali P, Avadhanula V, Santos HH, Weesner K, Hopkins L, Piedra PA, Maresso AW, Stadler LB. 2021. Evaluating recovery, cost, and throughput of different concentration methods for SARS-CoV-2 wastewater-based epidemiology. Water Research 197:117043. DOI: 10.1016/j.watres.2021.117043.

Natarajan A, Zlitni S, Brooks EF, Vance SE, Dahlen A, Hedlin H, Park RM, Han A, Schmidtke DT, Verma R, Jacobson KB, Parsonnet J, Bonilla HF, Singh U, Pinsky BA, Andrews JR, Jagannathan P, Bhatt AS. 2022. Gastrointestinal symptoms and fecal shedding of SARS-CoV-2 RNA suggest prolonged gastrointestinal infection. Med 3:371-387.e9. DOI: 10.1016/j.medj.2022.04.001.

Peccia J, Zulli A, Brackney DE, Grubaugh ND, Kaplan EH, Casanovas-Massana A, Ko AI, Malik AA, Wang D, Wang M, Warren JL, Weinberger DM, Arnold W, Omer SB. 2020. Measurement of SARS-CoV-2 RNA in wastewater tracks community infection dynamics. Nature Biotechnology 38:1164–1167. DOI: 10.1038/s41587-020-0684-z.

Prado T, Fumian TM, Mannarino CF, Resende PC, Motta FC, Eppinghaus ALF, Chagas do Vale VH, Braz RMS, de Andrade J da SR, Maranhão AG, Miagostovich MP. 2021. Wastewater-based epidemiology as a useful tool to track SARS-CoV-2 and support public health policies at municipal level in Brazil. Water Research 191:116810. DOI: 10.1016/j.watres.2021.116810.

Rieckermann J, Neumann M, Ort C, Huisman JL, Gujer W. 2005. Dispersion coefficients of sewers from tracer experiments. Water Science and Technology 52:123–133. DOI: 10.2166/wst.2005.0124.

Roldan-Hernandez L, Graham KE, Duong D, Boehm AB. 2022. Persistence of Endogenous SARS-CoV-2 and Pepper Mild Mottle Virus RNA in Wastewater-Settled Solids. ACS ES&T Water:acsestwater.2c00003. DOI: 10.1021/acsestwater.2c00003.

Schang C, Crosbie ND, Nolan M, Poon R, Wang M, Jex A, John N, Baker L, Scales P, Schmidt J, Thorley BR, Hill K, Zamyadi A, Tseng C-W, Henry R, Kolotelo P, Langeveld J, Schilperoort R, Shi B, Einsiedel S, Thomas M, Black J, Wilson S, McCarthy DT. 2021. Passive Sampling of SARS-CoV-2 for Wastewater Surveillance. Environmental Science & Technology 55:10432–10441. DOI: 10.1021/acs.est.1c01530.

Schwarzenbach RP, Gschwend PM, Imboden DM. 2017. Environmental organic chemistry. Hoboken, N.J: Wiley.

United States Environmental Protection Agency. 1999. Method 160.5, Methods and Guidance for the Analysis of Water Version 2.0.

Wade MJ, Lo Jacomo A, Armenise E, Brown MR, Bunce JT, Cameron GJ, Fang Z, Farkas K, Gilpin DF, Graham DW, Grimsley JMS, Hart A, Hoffmann T, Jackson KJ, Jones DL, Lilley CJ, McGrath JW, McKinley JM, McSparron C, Nejad BF, Morvan M, Quintela-Baluja M, Roberts AMI, Singer AC, Souque C, Speight VL, Sweetapple C, Walker D, Watts G, Weightman A, Kasprzyk-Hordern B. 2022. Understanding and managing uncertainty and variability for wastewater monitoring beyond the pandemic: Lessons learned from the United Kingdom national COVID-19 surveillance programmes. Journal of Hazardous Materials 424:127456. DOI: 10.1016/j.jhazmat.2021.127456.

Wolfe MK, Archana A, Catoe D, Coffman MM, Dorevich S, Graham KE, Kim S, Grijalva LM, Roldan-Hernandez L, Silverman AI, Sinnott-Armstrong N, Vugia DJ, Yu AT, Zambrana W, Wigginton KR, Boehm AB. 2021a. Scaling of SARS-CoV-2 RNA in Settled Solids from Multiple Wastewater Treatment Plants to Compare Incidence Rates of Laboratory-Confirmed COVID-19 in Their Sewersheds. Environmental Science & Technology Letters 8:398–404. DOI: 10.1021/acs.estlett.1c00184.

Wolfe MK, Topol A, Knudson A, Simpson A, White B, Vugia DJ, Yu AT, Li L, Balliet M, Stoddard P, Han GS, Wigginton KR, Boehm AB. 2021b. High-Frequency, High-Throughput Quantification of SARS-CoV-2 RNA in Wastewater Settled Solids at Eight Publicly Owned Treatment Works in Northern California Shows Strong Association with COVID-19 Incidence. mSystems 6. DOI: 10.1128/mSystems.00829-21.

Yang S, Dong Q, Li S, Cheng Z, Kang X, Ren D, Xu C, Zhou X, Liang P, Sun L, Zhao J, Jiao Y, Han T, Liu Y, Qian Y, Liu Y, Huang X, Qu J. 2022. Persistence of SARS-CoV-2 RNA in wastewater after the end of the COVID-19 epidemics. Journal of Hazardous Materials 429:128358. DOI: 10.1016/j.jhazmat.2022.128358.

Ye Y, Ellenberg RM, Graham KE, Wigginton KR. 2016. Survivability, Partitioning, and Recovery of Enveloped Viruses in Untreated Municipal Wastewater. Environmental Science & Technology 50:5077–5085. DOI: 10.1021/acs.est.6b00876.

Yin Z, Voice TC, Tarabara VV, Xagoraraki I. 2018. Sorption of Human Adenovirus to Wastewater Solids. Journal of Environmental Engineering 144:06018008. DOI: 10.1061/(ASCE)EE.1943-7870.0001463.

Zambrana W, Catoe D, Coffman MM, Kim S, Anand A, Solis D, Sahoo MK, Pinsky BA, Bhatt AS, Boehm AB, Wolfe MK. 2022. SARS-CoV-2 RNA and N Antigen Quantification via Wastewater at the Campus Level, Building Cluster Level, and Individual-Building Level. ACS ES&T Water:acsestwater.2c00050. DOI: 10.1021/acsestwater.2c00050.

