## Supporting Information for "Wastewater monitoring of SARS-CoV-2 RNA at K-12 schools: Comparison to pooled clinical testing data"

**Dimensional analysis**

*Solids*

In order to convert from X copies/uL from ddPCR to Y copies/g dry weight, the following equation was used for all samples.

$$\boldsymbol{X}\frac{\boldsymbol{copies}}{\mu L\ rxn}\times\frac{\boldsymbol{Volume\ of\ rxn}\ (\mu L)}{\boldsymbol{Volume\ of\ template\ in\ rxn}\ (\mu L)}\times\boldsymbol{dilution\ factor}$$
$$\times\frac{\boldsymbol{Volume\ of\ eluent\ from\ extract}\ (\mu L)}{\boldsymbol{Wet\ mass\ of\ solids\ in\ extract}\ (\boldsymbol{g})}$$
$$\times\ \boldsymbol{\%\ solids\ of\ sample}\ =\ \boldsymbol{Y}\frac{\boldsymbol{copies}}{\boldsymbol{g\ dry\ weight}}$$

*Liquids*

In order to convert form X copies/uL from ddPCR to Y copies/mL wastewater, the following equation was used for all samples.

$$\boldsymbol{X}\frac{\boldsymbol{copies}}{\mu L\ rxn}\times\frac{\boldsymbol{Volume\ of\ rxn}\ (\mu L)}{\boldsymbol{Volume\ of\ template\ in\ rxn}\ (\mu L)}\times\boldsymbol{dilution\ factor}$$
$$\times\frac{\boldsymbol{Volume\ of\ eluent\ from\ extract}\ (\mu L)}{\boldsymbol{Volume\ filtered}\ (\boldsymbol{mL})}$$
$$=\boldsymbol{Y}\ \frac{\boldsymbol{copies}}{\boldsymbol{mL\ wastewater}}$$

**Supplementary wastewater data methods**

Here, we briefly describe the methods used by the regional monitoring program to obtain RNA concentrations of Pepper Mild Mottle Virus (PMMoV) and SARS-CoV-2 N and S genes from settled solids at the wastewater treatment plant that processes the sewage from the sewershed that School A and B were part of. Wastewater primary sludge samples were collected from the primary settling tank of the wastewater treatment plant. Samples were dewatered by centrifuging. A small mass was suspended in DNA/RNA Shield (Zymo Research, California) spiked with a known concentration of bovine coronavirus vaccine (BCoV, PBS Animal Health, Ohio, Calf-Guard Cattle Vaccine), which acted as an internal control to ensure extraction recovery. The solution was homogenized by bead beating using 5/32” stainless steel grinder balls (OPS Diagnostics, New Jersey) and then centrifuged. The supernatant was subjected to nucleic acid extraction in 10 replicate aliquots using Chemagic Viral DNA/RNA 300 kit H96 (Perkin-Elmer, Massachusetts). Subsequently Zymo OneStep-96 PCR Inhibitor Removal Kit was used to remove inhibitors. The nucleic acids were used as templates in ddRT-PCR to measure the N and S gene of SARS-CoV-2. Each extraction replicate was quantified in its own well, making 10 replicate wells for each sample. For BCoV, the spiked-in internal recovery control, and PMMoV, a fecal strength indicator and an endogenous internal recovery control, were diluted 1:100 in water and run in 10 replicate wells. Extraction negative controls, extraction positive controls, no-template controls (NTCs), and PCR positive controls were run on each plate. PCR positive controls for SARS-CoV-2, BCoV, and PMMoV consisted of the following: guide RNA of target gene in SARS-CoV-2 (ATCC VR-1986D) and double stranded DNA gene blocks of BCoV and PMMoV, respectively. Detailed protocols by Topol et al., including specific concentrations and sequences of primers and probes, can be found on protocols.io.

The sewershed level data used for this study was the resulting RNA concentrations of N gene and S gene of SARS-CoV-2 along with PMMoV obtained from the solid primary sludge sample ranging from April 1, 2022 to June 10, 2022 in cp/g dry weight. The data can be found in the Stanford Data Repository (https://purl.stanford.edu/km945rd8103).

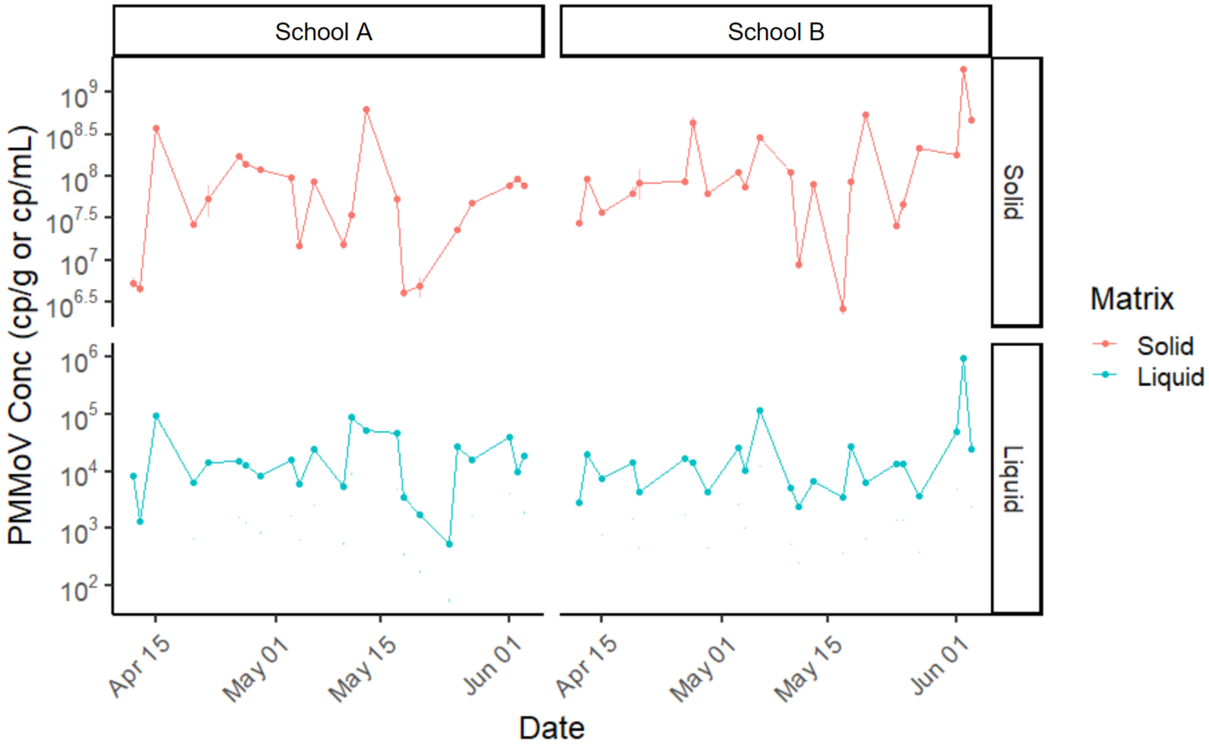


**Figure S1**. **PMMoV RNA concentration for each of the schools.**
Concentration in solid samples in units of cp/g dry weight (top) and concentration in liquid samples in units of cp/mL of wastewater (bottom). Error bars show the standard deviation as "total error" from ddPCR. Note that some error bars may not be visible when the associated standard deviation is very small.

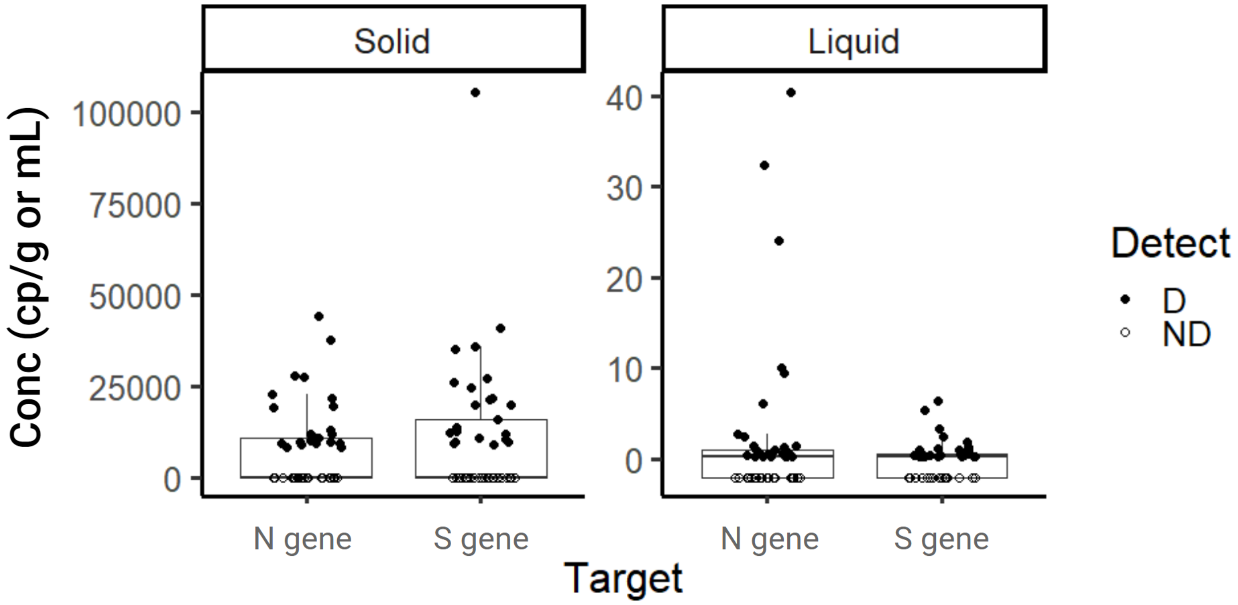


**Figure S2. Concentration distribution for targets N and S of SARS-CoV-2.**
Samples above the lower measurement limit are shown as filled circles. Samples that resulted in ND, shown as empty circles, were substituted with values slightly below zero to aid visualization.

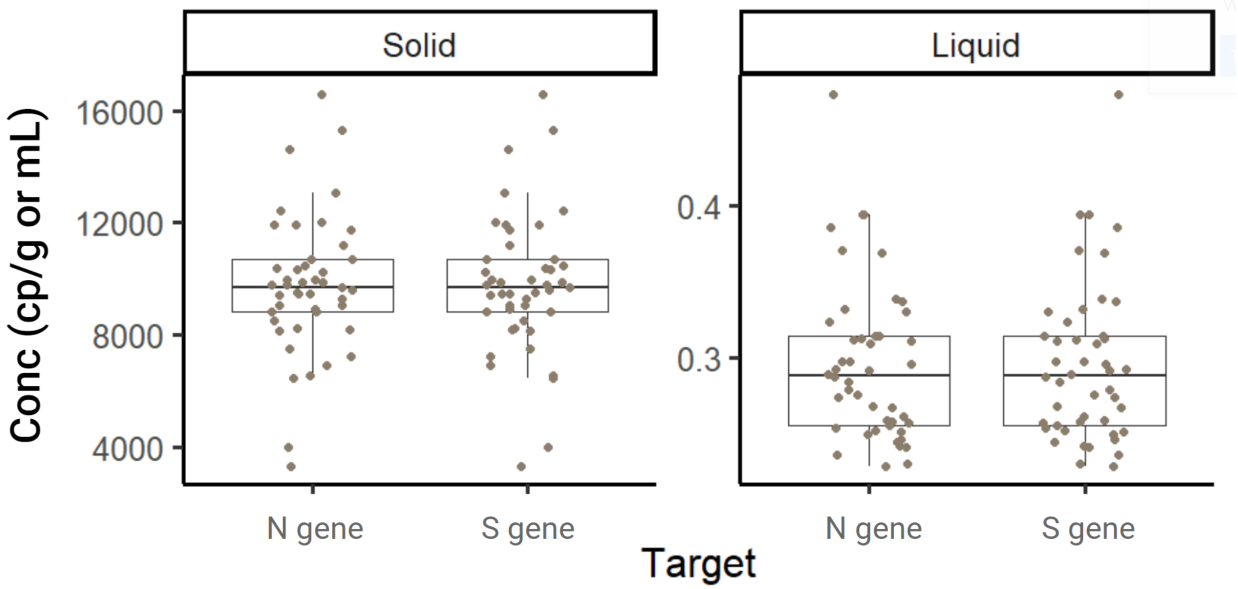


**Figure S3. Lowest theoretical measurement limit calculated for all samples analyzed in this study.**

The lowest theoretical measurement limit differs for each sample because 1) different number of droplets were generated for each sample and 2) associated fraction of dry mass or filtered wastewater differs slightly from sample to sample. The following formula was used to calculate the lowest theoretical measurement limit and then converted to appropriate units subsequently:

$$Concentration(cp/uL) = -ln(\frac{total\ accepted\ droplets - 3}{total\ accepted\ droplets}) \ / \ 0.00085uL\ .$$

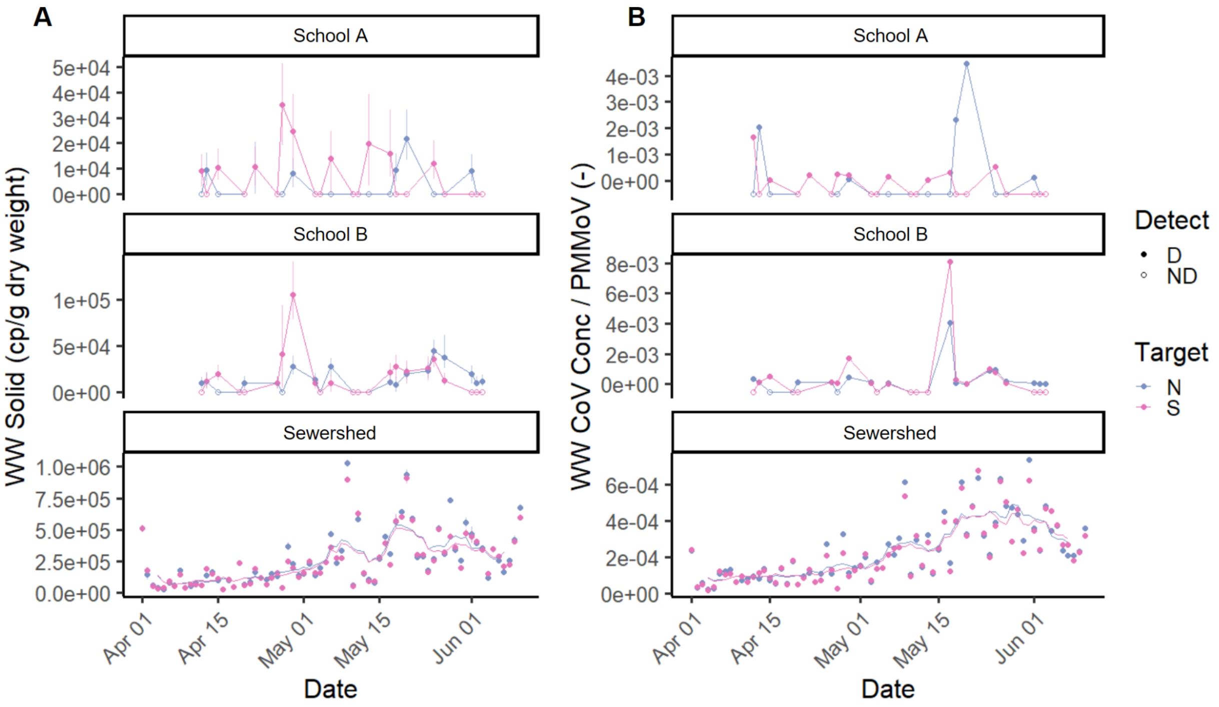


**Figure S4. Comparison of SARS-CoV-2 RNA concentration from schools to sewershed.** Data for only solid samples shown as the sewershed level data is of solid samples. Concentration in cp/g dry weight (A), and concentration normalized by PMMoV (B) are shown.

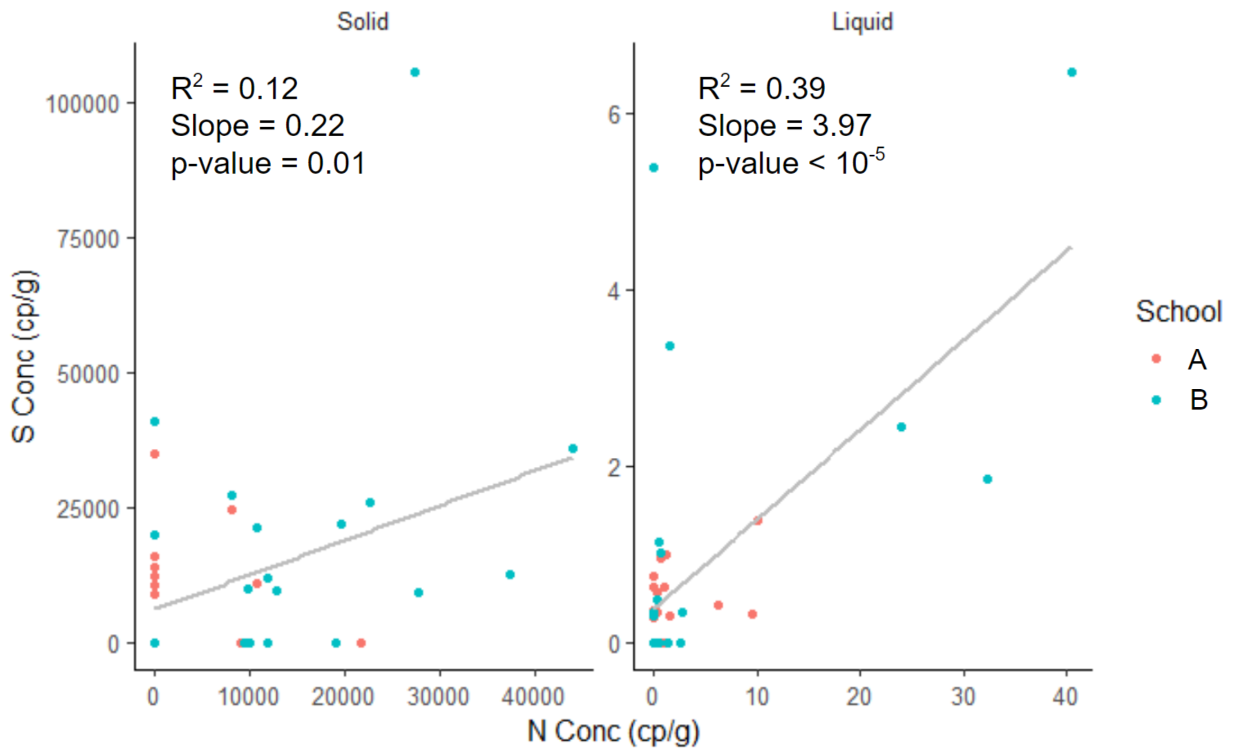


**Figure S5. Concentration of N gene and S gene in each sample.**
Zero was substituted for non-detects. Each data point represents SARS-CoV-2 RNA concentration from a single sample. The line shows the result of a linear regression between N gene and S gene RNA concentrations. The $R^2$ value, slope, and p-value are noted on each plot.

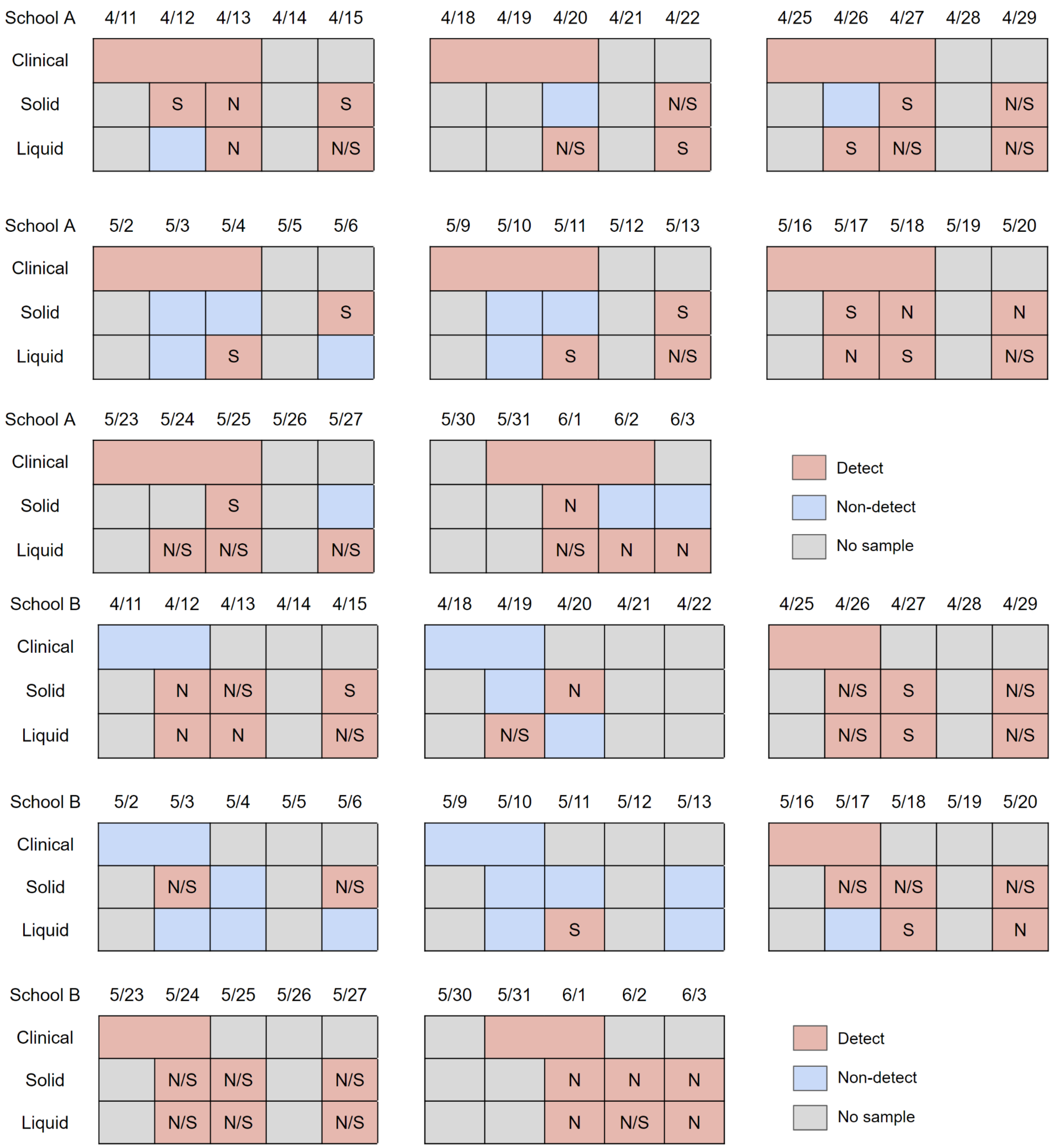


**Figure S6. Detection/non-detection compared between solid and liquid fraction of wastewater, and clinical pooled testing results.**

Clinical pooled testing was shown as complete sets as merged cells without distinguishing the cohorts tested on different days in order to show the testing result for the whole school. For wastewater samples, the detected gene target (N or S gene of SARS-CoV-2) is written on each day.

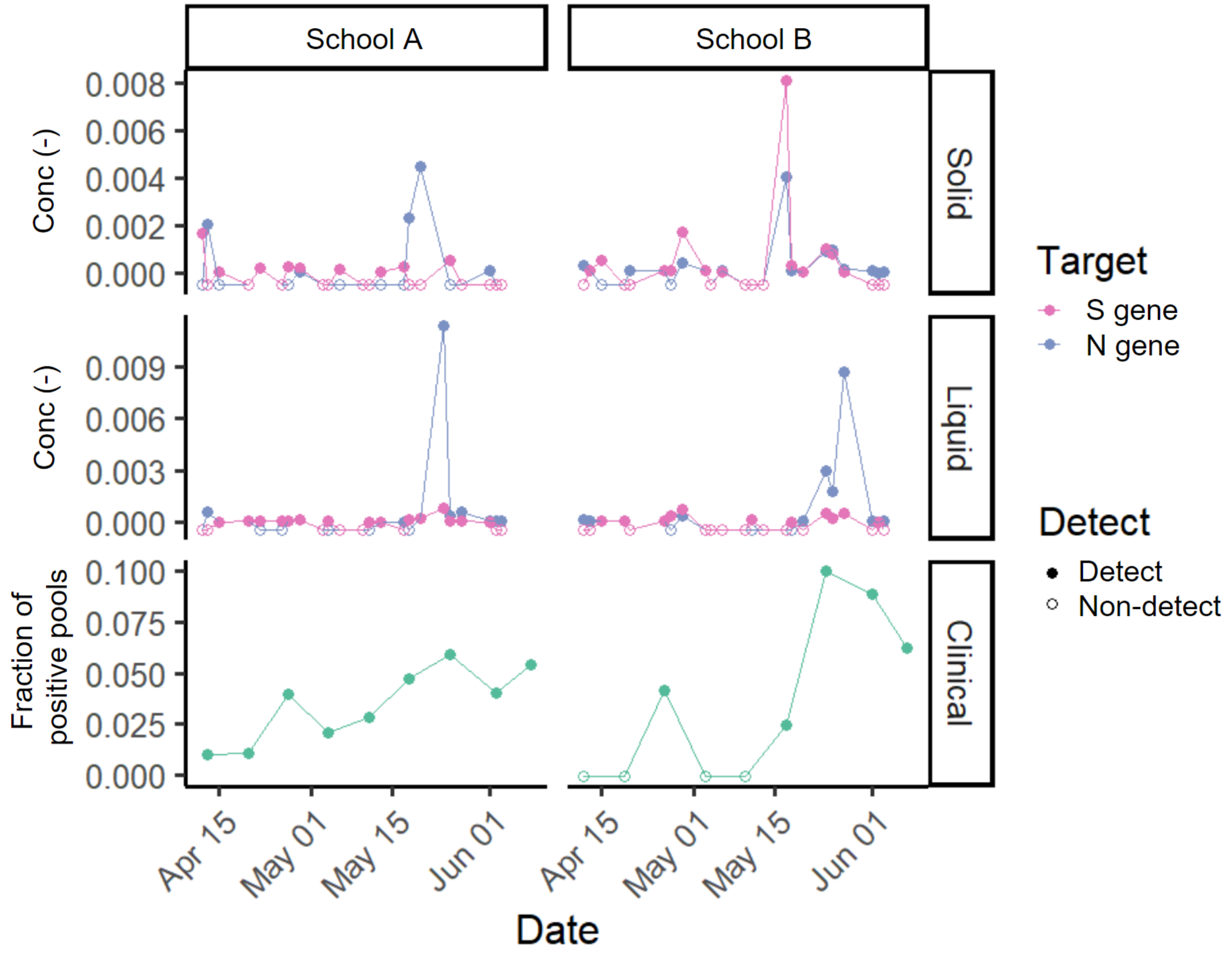


**Figure S7. Time series of SARS-CoV-2 measured normalized by PMMoV.**
SARS-CoV-2 targets N or S measured in solid samples (top) and liquid samples (middle) normalized by PMMoV, and fraction of positive pools for each of the schools (bottom) in the study over eight weeks. Each wastewater data point represents SARS-CoV-2 RNA concentration for a single sample. Samples below the lower measurement limit are shown as empty circles at negative value to aid with visualization. For clinical samples, empty circles represent no positive pools.

**Table S1. Association between SARS-CoV-2 RNA concentration from school wastewater to wastewater samples of the wastewater treatment plant that treats the sewershed that both schools are part of.**

1000 instances of Kendall's tau were calculated by sampling between upper and lower confidence intervals of each measured concentration of SARS-CoV-2 RNA. Empirical p-value was calculated as a percentage of Kendall's tau that resulted in a negative value.

| | **School A** | | **School B** | |
|---|---|---|---|---|
| | **Kendall's tau** | **p-value** | **Kendall's tau** | **p-value** |
| **N** | 0.08 | 0.25 | 0.23 | 0 |
| **S** | -0.09 | 0.81 | 0.11 | 0.047 |
| **N/PMMoV** | 0.10 | 0.14 | 0.04 | 0.18 |
| **S/PMMoV** | 0.004 | 0.53 | -0.06 | 0.86 |
